# Complementing Behavioral Weight Loss with Breath-based Yoga: A Proof-of-Concept and Initial Feasibility Study

**DOI:** 10.64898/2026.07.27.26358493

**Authors:** Sarah A. Purcell, Paul Dallaghan, Victoria Catenacci, Kristen Bing, Kaja Falkenhain, Ann E. Caldwell

**Affiliations:** Department of Medicine, Southern Medical Program, Centre for Chronic Disease Prevention and Management, University of British Columbia, Kelowna, British Columbia, Canada; School of Health and Exercise Sciences, University of British Columbia - Okanagan, Kelowna, BC, V1V 1V7 Canada; School of Medicine, Division of Endocrinology Metabolism and Diabetes, University of Colorado - Anschutz Medical Campus, Aurora, CO, 80045, USA; Anschutz Health and Wellness Center, University of Colorado - Anschutz Medical Campus, Aurora, CO, 80045, USA; Kaivalyadhama Yoga Institute, Lonalvala, India; Pennington Biomedical Research Center, Louisiana State University, Baton Rouge, LA, USA

**Keywords:** Mind-body therapies, health behavior, obesity management, breath-based yoga, complementary and alternative medicine, ORBIT model

## Abstract

**Background:** Yoga may support changes in dietary intake and/or physical activity levels, supporting sustainable weight loss. However, previous research has focused on yoga postures (*asanas*) rather than breathing practices (*pranayama*). This study used the proof-of-concept stage in the ORBIT model to evaluate feasibility, acceptability, and preliminary efficacy of a breath-based yoga program integrated within a behavioral weight loss (BWL) intervention.

**Methods:** Fifteen participants with overweight or obesity (BMI: 31.3±4.3 kg/m², 80% female) were enrolled in a 16-week, single-arm, group-based BWL intervention which included four breath-based yoga sessions/week, individualized daily goals for energy intake reduction, and progressive aerobic exercise recommendations. Primary outcomes included prespecified benchmarks enrollment, retention, adherence, and acceptability. Secondary outcomes included changes in body weight and composition, metabolic health indices (e.g., blood pressure, fasting glucose, abdominal obesity), physical activity, appetite, and dietary intake.

**Results:** Mean percent weight loss was 6.5 ± 6.0%, exceeding the pre-defined proof-of-concept benchmark. Recruitment and adherence exceeded established benchmarks, with mean in-class attendance of 84.1±14.6% and adherence to the prescribed duration of at-home yoga sessions averaging 85.5±38.4%. However, retention was lower than the pre-specified benchmark (73% vs 80%). Positive changes were observed in waist circumference, blood pressure, and appetite traits. Self-reported satisfaction with the program and outcomes were acceptable (mean ratings >6/10).

**Conclusions:** A BWL with a breath-based, low-intensity yoga can lead to acceptable weight loss with potential benefits in key behavioral and metabolic health indices. Recruitment, high adherence, and satisfaction demonstrated feasibility.

## 1. Introduction

Despite significant efforts to reverse the high prevalence of overweight and obesity, rates have continued to increase over the past two decades [1] resulting in substantial economic, medical, and quality of life burden [2]. Although anti-obesity medications and metabolic bariatric surgery are increasingly popular forms of obesity treatment, the first line of treatment recommended in current obesity guidelines is lifestyle modification or behavioral weight loss approaches (BWL) consisting of a reduced calorie diet, increased physical activity (PA), and behavioral support [3]. This behavior modification approach to treat obesity and reduce the risk of comorbid conditions often produces clinically meaningful weight loss of 5-10% in the short-term (∼6 months); however, about 50% of lost weight is typically regained within one year and long-term success in maintaining weight loss is poor [4]. It is critical to rigorously develop and evaluate adjunctive strategies that produce more widespread and sustainable improvements in dietary intake, PA, and weight loss and subsequent maintenance to reduce chronic disease risk.

There has been a growing interest in examining the effects of yoga on weight loss, weight loss maintenance, and cardiometabolic disease risk among adults with obesity and comorbid conditions such as metabolic syndrome, pre-diabetes, and diabetes [5–8]. Preliminary evidence has demonstrated that yoga may improve cardiometabolic disease risk both directly and indirectly through alterations in stress and stress reactivity, cognitive processes that support health behaviors (e.g., self-regulation and executive function), psychological well-being, sleep, physical mobility/pain, and reductions in food cravings and hunger [9–11]. Results are promising, but narrative and meta-analytical reviews of the literature note that more high-quality studies are needed [5,12,13]. Moreover, yoga is often broadly defined and variably implemented in existing research, and typically reflects the way yoga has primarily been adopted as a form of exercise in the modern world. However, traditional yoga encompasses body-, breath-, and mind-based practices that synergistically reinforce one another. To evaluate yoga’s potential as a complementary mind-body approach, yoga interventions should be designed with reference to their historical base and potential benefits of specific practices with clear descriptions of yoga practices to improve scientific rigor and reproducibility. While existing studies demonstrate movement-based yoga interventions are feasible in adults with overweight or obesity [14], the acceptability of a comprehensive, body-breath- and mind-based yoga intervention and effects of such on chronic disease risk merits investigation.

Proof-of-concept studies can provide a structured approach to preliminarily evaluate the integration of yoga within BWL. The primary goal of proof-of-concept studies within the ORBIT model for behavioral treatment development is to assess whether a fixed intervention can produce a clinically meaningful improvement in a primary outcome, using a quasi-experimental design in a small sample. The focus is on evaluating the intervention itself, with success determined by clinical rather than statistical significance, and the findings informing whether the intervention warrants further evaluation in a rigorously designed randomized trial [15]. Based on prior literature proposing hypothesized mechanisms through which yoga may influence obesity [8] (Phase Ia of the ORBIT model [16]) and evidence supporting its safety and potential benefits in individuals with obesity (Phase Ib) [6], we conducted a Phase IIa proof-of-concept study, consistent with the ORBIT model framework, to identify clinically meaningful signals in behavioral and physiological processes relevant to weight management. As such, the primary aim of this single-arm cohort intervention was to evaluate feasibility and acceptability of a BWL + breath-based yoga intervention in adults with obesity. The secondary aims were to evaluate weight changes using pre-defined benchmarks, and to explore changes in body composition, energy expenditure, cardiometabolic health markers, and behaviors that are important for weight loss, maintenance, and reducing cardiometabolic disease risk.

## 2. Methods

### 2.1 Participants

Volunteers from the greater Denver and Aurora metro areas were recruited to participate in the study using email announcements, community flyers, and posting through local social media groups from August 5, 2021 - September 27, 2021. All study procedures were approved by the Colorado Multiple Institutional Review Board and were performed at the University of Colorado – Anschutz Medical Campus. All participants provided written informed consent prior to data collection. This study was registered at clinicaltrials.gov (ID#: NCT05031221).

Interested individuals completed an online pre-screening questionnaire to determine potential eligibility. Participants underwent a physical examination by a medical provider including a health history form, blood draw, and resting electrocardiogram to determine eligibility. Primary inclusion criteria were age: 18-55 years; body mass index (BMI) 27-45 kg/m^2^, not regularly practicing yoga over previous 6 months; sedentary (<150 minutes per week of habitual moderate or higher intensity PA); and living or working within 30 minutes of the University of Colorado – Anschutz Medical Campus. Primary exclusion criteria were any physical or medical conditions that would preclude safely engaging in the interventions (i.e., cardiovascular disease, cancer, musculoskeletal, neurological, and psychiatric disorders) or abnormal values of fasting glucose (≥126 mg/dL), glycated hemoglobin (HbA1c; ≥6.5%), thyroid stimulating hormone (<0.4 or >4 mIU/L), hematocrit (<42% or >50% for males and <36% or >48% for females), white blood cell count (<4.5 or >11.0 × 10^9^/L) or platelets (<50,000 or >450,000 platelets/mcl), triglycerides (>400 mg/dL), low-density lipoprotein (LDL) cholesterol (>200 mg/dL), or an abnormal electrocardiogram. Participants were also excluded for nicotine use, drug or alcohol abuse, taking medications known to affect appetite, metabolism, or body weight; prior weight loss surgery; eating disorders; and weight change > 5% in the prior 3 months. Females who were currently pregnant, lactating, or pregnant within the prior 6 months were also excluded.

### 2.2 Intervention

#### 2.2.1 Traditional Yoga Practice Training Program

The yoga intervention consisted of twelve guided instructional yoga videos ranging from 20-42 minutes. Videos were designed and led by co-author PD (who has over 25 years of advanced yoga study and teaching experience with globally recognized Indian experts, Sri O.P Tiwari and Patthabi Jois). For the first 12 weeks, participants completed each video once in-person in a group setting (supervised by AEC and/or SAP) following the BWL class in the Anschutz Health and Wellness Center. Participants were then asked to complete the weekly video 3 additional times that week at home on their own, using a secure cloud-based platform (Wistia; Boston, MA, USA). All equipment needed (i.e., yoga mats, blocks) to complete the yoga sessions was provided. In weeks 1-12, participants were provided with a new video each week; in weeks 13-16, there were no in-person classes and participants were asked to complete videos from weeks 9-12 once/week in any order. Yoga sessions were designed to accommodate the anatomy and range of motion limitations for individuals with high BMI. The sessions were built with the goal of remaining true to the historical principles and original intent of ancient yogic teachings, while incorporating scientific justification for modern-world applications and delivery of classical practices from Patanjala and Hatha yoga. Each session started with brief Surya Namaskar Vinyasa (sun salute-based sequence) linking posture, slow movement, and breath. The majority of each session consisted of *pranayama* breathwork practices and inner focus meditation techniques. All components emphasized improving self-awareness.

#### 2.2.2 Behavioral Weight Loss Intervention

Participants received a comprehensive 16-week group-based BWL program adapted for this study from the *D*aily Caloric *R*estriction vs. *I*ntermittent *F*asting *T*rial (DRIFT) daily caloric restriction weight loss curriculum [17] which was based on obesity treatment guideline recommendations [3]. Weekly group meetings were taught by an experienced registered dietitian using a skills-based approach with cognitive and behavioral strategies for lifestyle modification [18,19]. Weekly meetings lasted approximately 60 minutes and took place in-person until no longer permitted due to the Omicron surge of the COVID pandemic, when classes were held over zoom in weeks 12 and 14. No class was held in week 13 during winter holidays, but participants were encouraged to keep up with the yoga and PA changes. Adherence to yoga and PA recommendations were tracked via Wistia and Fitbit for 16 weeks, and food logs were tracked over 15 weeks. BWL recommendations consisted of an individualized daily energy intake goal designed to produce a 30% energy deficit from weight maintenance requirements based on measured resting metabolic rate (RMR) via indirect calorimetry (RMR x activity factor of 1.5) [20], with a suggested macronutrient content of 55% carbohydrates, 15% protein, and 30% fat. The curriculum used a combination of large group discussion, small breakout discussions, visual demonstrations, and written exercises. Example topics included: realistic weight loss goal setting, self-monitoring strategies, cognitive restructuring, improving personal food environments and social networks, strategies to overcome barriers to healthy eating and increasing PA, incremental goal setting, and providing feedback on goals. Participants were not withdrawn for non-adherence.

Participants were also given a prescription to gradually increase their PA to 150 minutes/week in line with the diabetes prevention program recommendations for reducing diabetes risk [21] and the American College of Sports Medicine guidelines [22]. Participants were instructed in how to use a relative intensity scale [23] to achieve the target moderate intensity and were provided Fitbits (San Francisco, CA, USA) to track activity and intensity using the built-in heart rate monitor.

### 2.3 Feasibility and Acceptability

Primary outcomes were enrollment, retention, collection of clinical outcome measures, acceptability, adherence, and adverse events. An enrollment target of 12-20 eligible individuals over a 12-week recruitment window and attrition <20% over 16 weeks of intervention were determined *a priori* to demonstrate feasibility of recruitment and retention. Feasibility of performing outcome measures was determined *a priori* as having >90% of enrolled participants completing primary and secondary outcome measures at baseline and follow-up. Adherence to BWL group classes and in-person yoga sessions was calculated as the total number of attendances divided by the number of classes it was possible to attend (i.e., 16 and 12, respectively). Adherence to the at-home yoga sessions was assessed as the number of sessions (frequency), of the number of minutes that videos were watched, and the total number of minutes that participants completed of the yoga videos on Wistia – the latter two metrics were expressed as percentages (i.e., divided by the total prescribed minutes x 100). Instances of viewing duration less than 10 minutes were excluded. Other engagement metrics to evaluate intervention adherence included self-reported energy intake and physical activity. Considering potential inaccuracies in reported energy intake [26], we defined adherence to the diet intervention as the percentage of weeks in which food was logged for all seven days. Adherence to exercise targets was defined as the percentage of weeks in which participants met their prescribed exercise goals, measured via Fitbit.

At weeks 8 and 16, participants provided feedback on the previous 8 weeks of the intervention using a study-specific questionnaire that consisted of 10-point Likert scales (1 being unfavorable / negative responses and 10 being favorable / positive responses), multiple answer, and yes-no responses. Adverse events were collected and recorded.

### 2.4 Proof-of-Concept for Weight Loss

Given the central role of weight change in BWL interventions, we established a proof-of-concept threshold for weight loss of 5.3% ± 1%. This value reflects the mean weight loss within ±1% achieved in the daily caloric restriction arm of a large randomized trial (n=165) conducted at our center - the Daily Caloric Restriction vs Intermittent Fasting Trial (DRIFT; NCT03411356) - in a similar patient population [17,24]. This benchmark was chosen because the DRIFT weight loss curriculum for the daily caloric restriction arm served as the basis for the group-based intervention used in the current study and was delivered by the same registered dietitian [17,24]. Note, however, that this threshold was not included in the initial trial registration, as the data needed to establish it were still being collected at the time of registration.

In both DRIFT and the current BWL + breath-based yoga intervention, body weight was measured using a calibrated digital scale with participants in a fasted state and wearing a hospital gown. Outcome assessments were conducted at similar time points; follow-up weight assessments occurred at week 13 (± 2 weeks) in DRIFT and at calendar week 16 (class week 14 ± 2 weeks) in the current study. Among DRIFT daily caloric restriction participants with available follow-up data (n=73), the mean percent weight loss was 5.3 ± 3.7% (unpublished data). Based on this reference, the threshold for successful proof-of-concept for the current intervention was defined as a mean percent weight loss between 4.3% and ≥ 6.3% among participants with follow-up data. For the purposes of data analyses and interpretation, weight loss ≥5% of baseline weight was deemed “clinically meaningful”, in accordance with current obesity treatment guidelines and prior research indicating that this level of weight reduction is associated with improved metabolic outcomes [25].

### 2.5 Anthropometric, Metabolic, and Behavioral Outcomes

#### 2.5.1 Study Day Visits

Baseline and follow-up visits occurred in a two-week window before and after the intervention to assess secondary outcomes related to preliminary clinical effectiveness/efficacy. Participants were asked to arrive at the University of Colorado Hospital’s Clinical and Translational Research Center (CTRC) after an overnight fast of at least 12 hours and avoiding alcohol for at least 24 hours and vigorous exercise for at least 48 hours.

#### 2.5.2 Anthropometric and Body Composition Outcomes

Height was measured using a wall-mounted stadiometer. Body mass index was calculated by dividing body weight in kilograms by height in meters squared and categorized according to the World Health Organization cut points [27]. Waist circumference was measured at the umbilicus using a tape measure. Measurements for body weight and waist circumference were taken twice, and the average of the values was used. Fat mass (in kg) and fat-free mass (FFM; in kg; lean mass + bone mineral density) were collected using full body dual X-ray absorptiometry calibrated according to the manufacturers’ instructions (DXA; Hologic Delphi-W, Hologic Inc., Bedford, MA).

#### 2.5.3 Metabolic Outcomes

Diastolic and systolic blood pressure were obtained on the right side of the body after a five-minute rest using an automated blood pressure cuff (Omron Autocuff, Omron Healthcare Co. Ltd., Lake Forest, IL, USA); two measures were taken and the average of these was used. Participants completed a 2-hour oral glucose tolerance test to measure glucose and insulin metabolism. An intravenous catheter was placed on the antecubital vein inside of the participant’s arm opposite the elbow to obtain blood samples in the fasted state and every 30 minutes for two hours after consumption of a 75-gram glucose beverage. Glucose values were converted to SI units (mmol/L) by multiplying mg/dL values by 0.0555. Homeostatic model assessment for insulin resistance (HOMA-IR) index was calculated as:

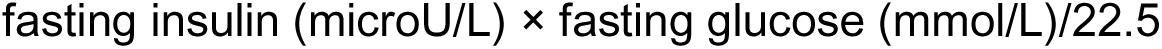

Glucose area under the curve was calculated according to the trapezoidal rule [28]. Glycated hemoglobin (HbA1c), total triglycerides, and total, high- and low-density lipoprotein cholesterol were also measured from the fasted blood sample. All samples were assessed by the CTRC lab core using the following techniques: hexokinase for glucose (Beckman Coulter; Brea, CA, USA; 0.6% within-day precision), chemiluminescent immunoassay for insulin (Beckman Coulter; 1.6% within-day precision), enzymatic assays for triglycerides and cholesterol (Beckman Coulter; 0.8% and 0.9% within-day accuracy, respectively) and potassium ferricyanide for HbA1c (Siemens; Munich, Germany; 1.8% within-day precision).

#### 2.5.4 Resting Metabolic Rate

RMR was measured using indirect calorimetry with a ventilated hood (TrueOne 2400 metabolic cart, Parvomedics, Sandy, UT, USA) to determine individual energy intake goals and characterize changes in energy expenditure across time. The flow meter and gas analyzers were calibrated before each test according to the manufacturer’s guidelines. Participants rested supinely in a quiet, thermoneutral (20-23°C), dimly lit room for 25-30 minutes before testing. Respiratory gas exchange was collected for 20-25 minutes, and the mean of the last 15 minutes of oxygen and carbon dioxide data were used to calculate RMR using the Weir equation [29]. RMR was expressed in absolute terms (kcal/day) and adjusted for body composition using a least squares best-fit linear regression equation with RMR as a function of the FFM and FM, similar to previous studies [30,31]. The resulting equation using baseline values was used to calculate adjusted RMR for each participant at baseline and follow-up, based on their corresponding FM and FFM values, as follows:

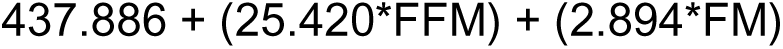

where FM and FFM were measured in kilograms. The difference between calculated and measured RMR was used to assess RMR unexplained by changes in body composition, termed ‘RMR residuals’. To fully account for the trajectory of change in body composition and energy expenditure, baseline residuals were subtracted from follow-up residuals [32]. Negative and positive RMR residuals indicated measured RMR values that were less than or greater than anticipated when accounting for body composition, respectively.

#### 2.5.5 Physical Activity and Sedentary Behavior

ActivPAL3^TM^ accelerometers (PAL Technologies, Glasgow, Scotland) were used to measure PA and sedentary behavior over seven consecutive days. The accelerometers were attached to the midline of the anterior thigh on participants’ dominant side, one-third of the way between the hip and the knee using waterproof Tegaderm. Data were downloaded and processed using the manufacturer’s software (PALBatch, V8) using the CREA enhanced analysis algorithm and a 10-hour wear time cutoff for validity. Behaviors were categorized according to automatic and manual calculations: total steps/day, total sedentary time, light PA (cadence <75 steps per minute), and moderate-to-vigorous PA (MVPA) (cadence ≥ 75 steps per minute).

#### 2.5.6 Dietary Intake and Appetite

Participants were asked to track their dietary intake over three days (two weekdays and one weekend day) using photographic food records by utilizing the camera function of their smartphones. Participants were instructed on the correct methods of recording dietary data, which included information on cooking method, serving size, added oils/sugar, and time of meal. Dietary data were entered and assessed using the Nutrition Data System for Research software by the CTRC staff as soon as possible after collection of food records. Energy intake was expressed in absolute terms (kcal/day) and adjusted for body weight (kcal/kg/day), and macronutrients were expressed as a percentage of energy intake. Only data which met the following parameters were included: 1) energy intake within each individual’s plausible range using the method proposed by McCrory *et al.*[33], and 2) at least three eating occasions (>100 kcal each) recorded that day.

During the three days of dietary intake collection, participants were also asked about their appetite each study day via text messages (sent using Twilio) containing links to a secure RedCap survey (‘appetite state’). Both appetite and fatigue were sampled at six specific time points each day (waking; +2, +6, +10, and +14 hours after waking, and at bedtime). Appetite-related questions asked participants about their current hunger, fullness/satiety, and prospective food consumption using a visual analog scale (0-100). Appetite scores were adjusted for previous mealtime as well as energy content collected via photographs of food.

The following aspects of appetite traits were assessed at baseline and follow-up: dietary restraint, disinhibition, hunger (Three Factor Eating Questionnaire [34]), appetite for palatable foods (Power of Food Scale [35]), and food cravings (Food Craving Inventory [36]). Questionnaires were scored according to the original publication’s guidelines, with greater scores indicating higher levels of the dimension.

### 2.6 Statistical Analyses

Data were analyzed using SPSS (version 29.0.1.0; IBM Corporation, Armonk, NY, USA) and R (version 4.3.2; R Foundation for Statistical Computing, Vienna, Austria). Continuous data are presented as mean ± standard deviation or median [interquartile range; IQR] and categorical data are presented as number (%). Normality was assessed using the Shapiro-Wilk test. Adherence and acceptability data were reported in both completers and non-completers to gain an understanding of feasibility and refinement for future studies. Given the purpose of a proof-of concept study is to identify clinically significant signals and test fixed protocols [16], we focused our analyses and interpretation of outcome measures on participants that completed the study (rather than an intention-to-treat approach) to evaluate the potential efficacy of the intervention under ideal conditions. This approach allowed us to assess outcomes among participants who adhered to the protocol, providing preliminary evidence to inform future, larger-scale trials. To measure changes in outcome variables, paired t-tests were performed in variables conforming to normality assumptions, while those with non-normal distribution were tested using Wilcoxon Rank Sum tests. Differences in age and BMI between participants who completed the intervention versus those who dropped out were compared using independent samples t-test or Mann-Whitney U-test. Statistical significance was established at α=0.05, but because this was a proof-of-concept and early feasibility study, we focused on effect sizes (Cohen’s *d*) rather than significance testing in interpretation of results. Effect sizes for changes in outcomes were interpreted as small (Cohen’s *d* < 0.2), medium (*d* = 0.2 – 0.5) and large (*d* > 0.5), and we focus the conclusions on outcomes with large effect sizes. To assess potential changes in appetite states, a linear mixed model including time point (baseline, follow-up) as a fixed effect, participant as a random effect, and adjustments for the day of the study, the time of the assessment, and energy content of the meal prior to the assessment. This approach was chosen for its flexibility in accommodating variability in the timing of meals and appetite ratings across participants. The findings should be considered as preliminary, given the study was not designed or powered to detect statistically significant changes across time.

## 3. Results

### 3.1 Participants and Feasibility

A total of 106 individuals expressed interest in the study over a 7-week recruitment window (**Figure 1**). Of those, 21 were eligible and consented to participate and 15 enrolled in the study, meeting the enrollment target. Eleven participants completed the 14-week intervention (73.3% retention). All participants completed outcome measures at baseline and 10 of 11 completed all 14-week outcome measures (90.9%). The sample was 80% female, 80% White, with high levels of education and household income, **Table 1**. Mean age of participants was 40 ± 10 years and mean BMI was 31.3 ± 4.3 kg/m^2^. There were no differences in age (completers: 39 ± 11; non-completers: 42 ± 8 years; p=0.662) or BMI at baseline (completers: 28.3 [27.0 – 33.8]; non-completers: 31.8 [30.9 – 37.8] kg/m^2^; p=0.086) between completers and non-completers.

**Figure 1.**
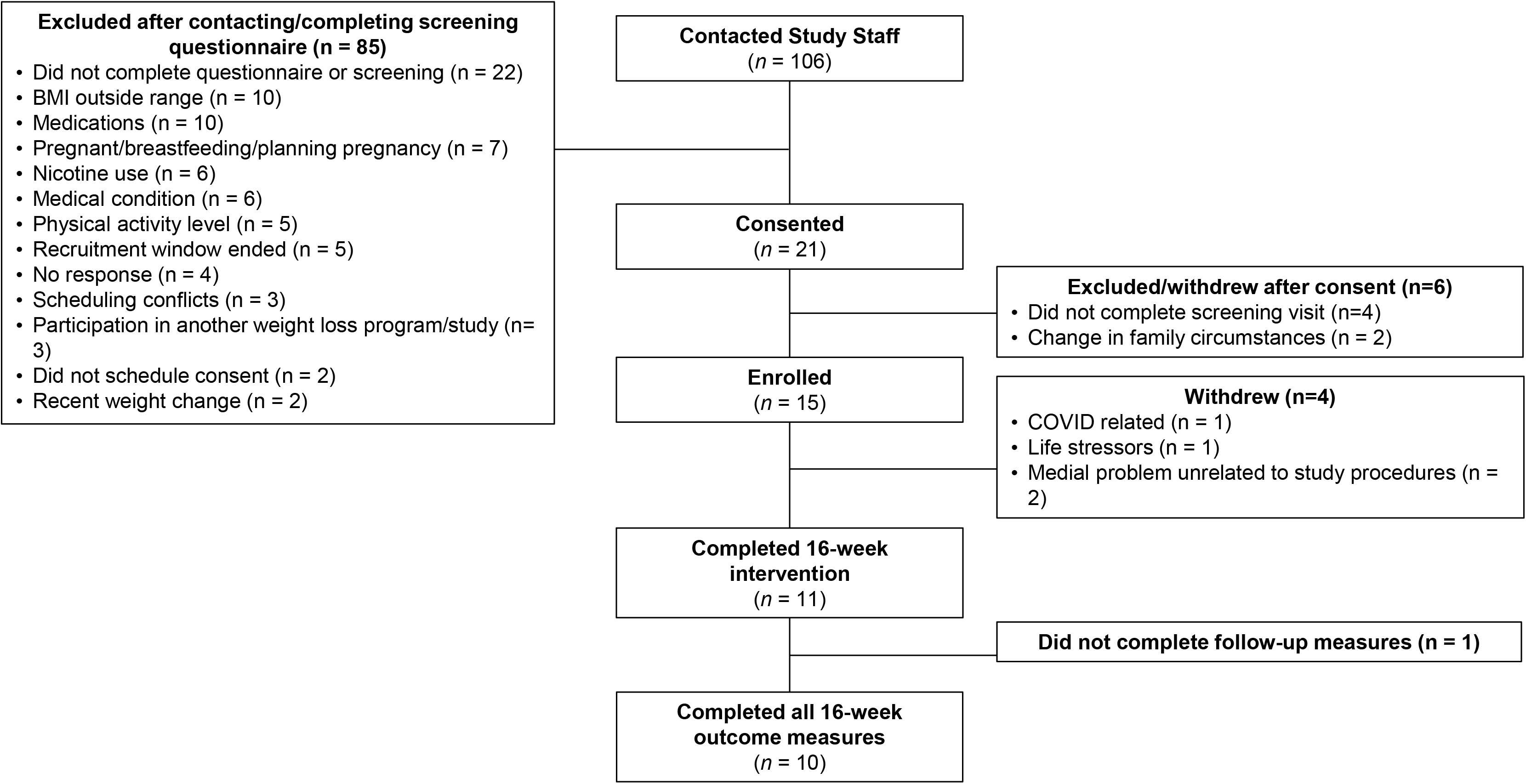
Participant flow for study inclusion.

**Table 1.** Sample characteristics at baseline.

| Characteristics | Mean ± SD or n (%) |
| --- | --- |
| Age, years | $40 \pm 10$ |
| BMI, kg/m<sup>2</sup> | $31.3 \pm 4.3$ |
| Sex, female | 12 (80) |
| Race |  |
| African American | 1 (6.7) |
| American Indian or Alaska Native | 1 (6.7) |
| Asian | 1 (6.7) |
| White | 12 (80.0) |
| Ethnicity |  |
| Hispanic or Latino | 3 (20) |
| Not Hispanic or Latino | 12 (80) |
| Education |  |
| 2-year college | 1 (6.7) |
| 4-year college | 6 (40.0) |
| Master's degree | 6 (40.0) |
| Doctorate | 2 (13.3) |
| Household income |  |
| 45-70 k/year | 4 (26.7) |
| 70-110 k/year | 3 (20.0) |
| >110 k/year | 6 (40.0) |
| prefer not to answer | 2 (13.3) |

N=15. BMI: body mass index

### 3.2 Proof-of-Concept

The intervention exceeded the pre-defined proof-of-concept benchmark. Participants who completed the intervention lost a mean of 6.5 ± 6.0% (median [IQR]: 4.6 [14.0, 1.9%] of initial body weight, equating to a mean absolute loss of 5.2 ± 4.9 kg. Weight loss was the result of a reduction in both FM and FFM. Five completers (45.0%) achieved clinically meaningful weight loss (≥5%) at 16 weeks, with three (27.3%) achieving ≥10% weight loss, **Figure 2**. Data were similar when only females were assessed (n=9; data not shown); hence data from both males (n=2) and females combined are presented.

**Figure 2.**
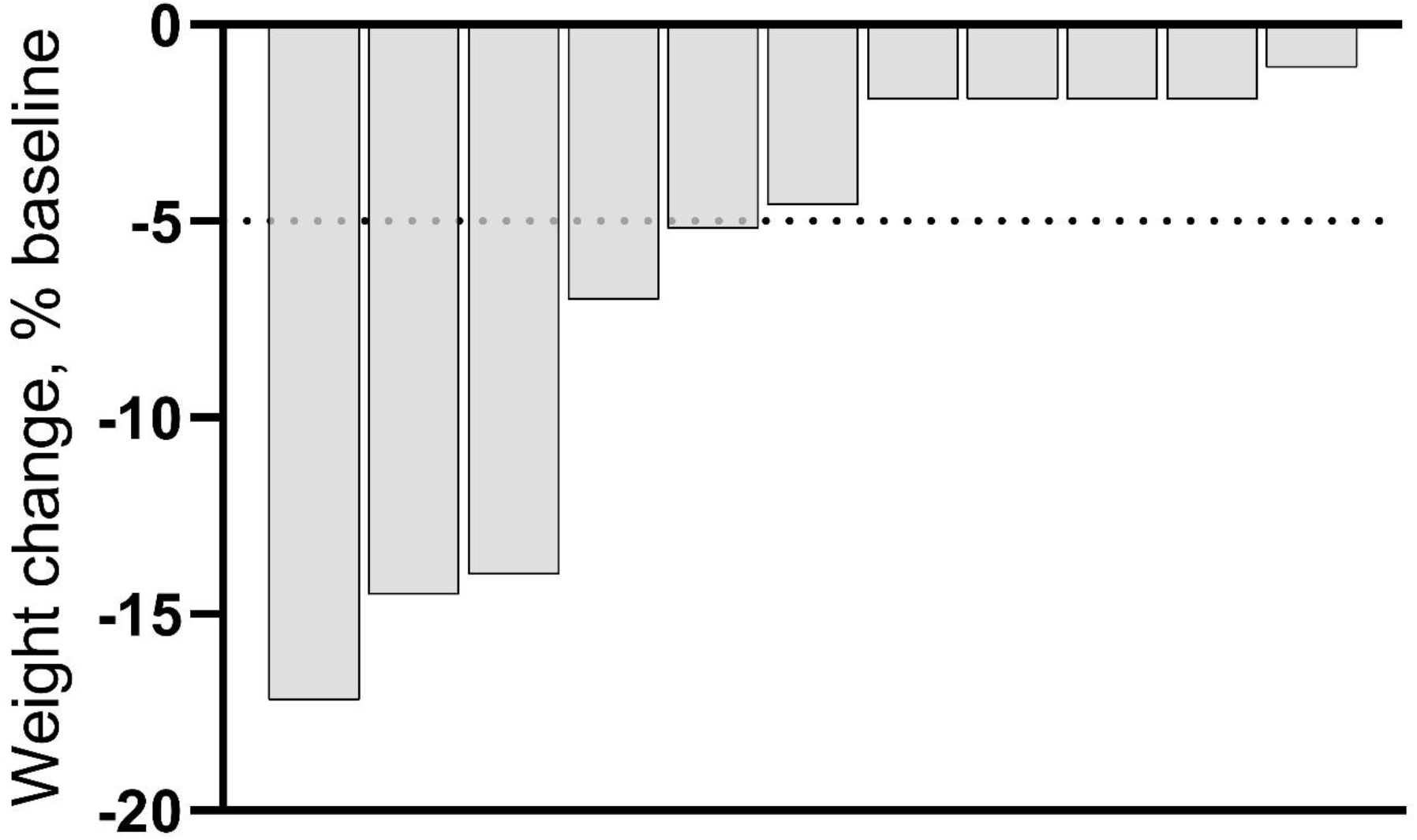
Waterfall plot of weight change during the intervention. Each bar represents an individual participant who completed the intervention.

### 3.3 Adherence and Acceptability

Group weight loss class attendance was 84.1 ± 14.6% (13.5 ± 2.3 classes) in completers and 69.2 ± 29.0% (11.1 ± 4.6 classes) in non-completers. Individuals who completed the intervention averaged 2.9 ± 1.3 at-home yoga sessions per week, corresponding to 91.8 ± 40.8% (median [IQR]: 100 [66.7 – 125.0]%) of the weekly target. Participants who discontinued the intervention completed 2.8 ± 1.4 at-home yoga sessions per week up to the point of drop out, which was 86.8 ± 42.7% (100 [66.7 - 100.0]%) of the weekly target. Among completers, the percent of video watched was 80.4 ± 28.4% (97.2 [70.4 – 99.9%]) and adherence to the duration of prescribed at-home yoga sessions was 85.5 ± 38.4% (99.1 [66.6 – 100.0%]) among completers. Among non-completers, the percent of video watched was 78.2 ± 30.3% (95.6 [66.8 – 99.9%]) and adherence to the at-home session duration was 80.6 ± 40.2% (96.1 [56.3 – 100.0%]). Among individuals completing the intervention, mean adherence to diet tracking was 77.3 ± 21.5% (12.4 ± 3.4 weeks) and mean adherence to exercise targets was 47.7 ± 21.3 % (7.6 ± 3.4 weeks). In non-completers, adherence to diet tracking was 62.1 ± 33.1% (9.9 ± 5.3 weeks) and adherence to exercise targets was 37.9 ± 25.2% (6.1 ± 4.0 weeks).

At intervention mid-point and completion, participants rated their ability to adhere to the program and overall satisfaction with the program and outcomes favorably (i.e., mean Likert scale ratings > 6), **Table 2**. Only completers provided feedback. Physical difficulty of the yoga sessions was rated low (week 8: 3.5 ± 1.8; week 16: 3.2 ± 1.9). Lack of time (selected by n=7 [63.6%]), vacation/travel, and being too busy (selected by n=6 [54.5%] each) were the primary barriers to adhering to the intervention. At week 8, approximately half of the sample indicated that stress (n=6 [54.5%]) and fatigue (n=5 [45.5%]) were major barriers to participation; fewer people indicated these were barriers at intervention completion (stress: n=4 [36.4%]; fatigue: n=2 [18.2%]). At intervention completion, the vast majority of participants (‘yes’ in n=10, 90.9% for all questions) indicated they observed positive changes in physical health (i.e., fatigue, strength, endurance, hunger, sleep quality/quantity, increased mobility), psychological health (i.e., stress, confidence, body satisfaction), overall well-being, and dietary intake as a result of the program, **Supplemental Table 1**.

**Table 2.** Ratings of yoga adherence and satisfaction.

|  | **Week 8** | **Week 16** |
| --- | --- | --- |
|  | **Mean ± SD** | **Mean ± SD** |
| Please rate your yoga adherence level during the past 8 weeks. | 8.3 ± 1.8 | 7.1 ± 2.3 |
| Please rate how hard it was to adhere to the prescribed yoga regimen during the past 8 weeks. | 5.5 ± 3.1 | 6.2 ± 2.7 |
| Please rate how likely it is that you can adhere to doing yoga when the study is over. | 8.8 ± 1.5 | 7.1 ± 3.2 |
| Please rate your satisfaction with this program with regard to ability to adhere to yoga as prescribed. | 7.8 ± 1.7 | 7.5 ± 1.9 |
| Please rate how much you have enjoying doing the yoga videos 4 times/week over the past 8 weeks. | 6.5 ± 2.2 | 6.5 ± 2.5 |
| Please rate how engaging the yoga practices have been. | 6.2 ± 2.1 | 5.8 ± 2.5 |
| Please rate your satisfaction with this program with regard to the content of the yoga intervention (including the weekly in-person sessions and corresponding videos). | 6.6 ± 2.0 | 6.9 ± 2.5 |
| Please rate your satisfaction with this program with regard to weight loss achieved to date. | 6.9 ± 2.5 | 6.7 ± 2.5 |
| Please rate your satisfaction with this program with regard to physical function (i.e., daily activities) achieved to date. | 7.4 ± 0.9 | 7.4 ± 2.2 |
| Please rate your satisfaction with this program with regard to improvements in fatigue levels. | 6.0 ± 2.2 | 6.6 ± 2.8 |

Scores are from a 1 – 10 scale with 1 being unfavorable or negative responses (e.g., ‘not adherent at all’) and 10 being favorable or positive responses (e.g., ‘perfect adherence’). n=11 at both timepoints (same individuals). One participant dropped out before week 11 and three did not complete the survey.

There were four anticipated adverse events that were probably or definitely related to the study protocol. All were mild in severity and related to performance of outcome measures rather than study intervention, and were expected with study procedures (two were due to skin irritation from the activity monitor, and two were associated with OGTT procedures). All were resolved without medical attention beyond the event.

### 3.4 Anthropometric, Metabolic, and Behavioral Outcomes

#### 3.4.1 Anthropometric and Metabolic Outcomes

The effect sizes for reductions in total body weight, BMI, and waist circumference were all large (Cohen’s *d* range, **Table 3**). The mean reduction in systolic blood pressure was particularly robust (from 122 ± 12 to 113 ± 11), evidenced by a large effect size (Cohen’s *d* = -0.81). Effect sizes for changes in the remaining indices of metabolic health were small to medium, but all changed in a direction suggestive of improved metabolic health. Total cholesterol, HbA1c, fasting insulin, fasting glucose, glucose area under the curve, and HOMA-IR index decreased with medium effect sizes.

**Table 3.** Changes in behavioral, metabolic, and physical outcomes.

|  | Baseline<br>N=15 | Follow-up<br>N=10 | Change<br>N=10 | Effect size<br>(Cohen's *d*) | p |
| --- | --- | --- | --- | --- | --- |
| **Anthropometrics & body composition** |  |  |  |  |  |
| Body weight, kg* | 87.5 ± 14.8 | 80.7 ± 17.8 | **-5.3 ± 4.9** | **-1.073** | 0.005 |
| Body mass index, kg/m<sup>2</sup>* | 31.3 ± 4.3 | 28.9 ± 5.6 | **-1.9 ± 1.7** | **-1.137** | 0.004 |
| Waist circumference, cm | 97.7 ± 11.4 | 89.3 ± 11.2 | **-8.3 ± 4.5** | **-1.842** | <0.001 |
| FM, kg | 34.9 ± 10 | 30.4 ± 11.5 | **-2.7 ± 4.6** | **-0.593** | 0.094 |
| FFM, kg | 51.7 ± 9.7 | 47.4 ± 9 | **-2.9 ± 1.2** | **-2.392** | < .001 |
| **Additional metabolic health indices** |  |  |  |  |  |
| Systolic blood pressure, mmHG | 122 ± 12 | 113 ± 11 | **-9 ± 10** | **-0.806** | 0.016 |
| Diastolic blood pressure, mmHG | 72 ± 8 | 71 ± 7 | 0 ± 7 | 0.052 | 0.868 |
| Triglycerides, mg/dL | 87 [59, 181] | 71 [60, 83] | -18 [-54, 18] | -0.097 | 0.878 |
| Cholesterol, mg/dL | 179 ± 45 | 173 ± 45 | -7 ± 24 | -0.311 | 0.351 |
| HDL cholesterol, mg/dL | 51 ± 17 | 51 ± 17 | 0 ± 3 | 0.035 | 0.915 |
| LDL cholesterol, mg/dL | 105 ± 36 | 103 ± 36 | -4 ± 21 | -0.187 | 0.568 |
| HbA1c, % | 5.4 ± 0.3 | 5.3 ± 0.3 | -0.1 ± 0.2 | -0.391 | 0.247 |
| Fasting insulin, μU/mL | 5.8 ± 3.1 | 4.7 ± 2.5 | -0.8 ± 1.9 | -0.414 | 0.223 |
| Fasting glucose, mmol/L | 4.9 ± 0.6 | 4.8 ± 0.6 | -0.1 ± 0.3 | -0.211 | 0.522 |
| Glucose AUC, mmol/L x 120 minutes | 768 [678, 927] | 726 [568, 883] | -17 [-111, 76] | -0.355 | 0.575 |
| HOMA-IR | 1.3 ± 0.8 | 1.0 ± 0.7 | -0.2 ± 0.4 | -0.405 | 0.232 |
| **Resting metabolic rate (RMR)** |  |  |  |  |  |
| Measured RMR, kcal/day | 1852 ± 297 | 1747 ± 227 | **-102 ± 170** | **-0.601** | 0.090 |
| Predicted RMR kcal/day | 1852 ± 252 | 1731 ± 242 | **-82 ± 37** | **-2.217** | <0.001 |
| RMR residuals, kcal/day | 0 ± 156 | -16 ± 116 | -20 ± 157 | -0.129 | 0.842 |
| **Dietary intake\*\*** |  |  |  |  |  |
| Energy intake, kcal/day | 1627 ± 317 | 1628 ± 356 | 20 ± 244 | 0.081 | 0.838 |
| Energy intake, kcal/kg body weight/day | 19.3 ± 3.4 | 21.7 ± 4.4 | 1.7 ± 3.3 | 0.117 | 0.766 |
| Fat, % total energy intake | 41.1 ± 6.4 | 40.4 ± 5 | -0.9 ± 7.8 | -0.076 | 0.847 |
| Carbohydrates, % total energy intake | 42.4 ± 6.9 | 41.9 ± 8.3 | -0.8 ± 10.3 | -0.175 | 0.660 |
| Protein, % total energy intake | 15.7 ± 2.8 | 17.5 ± 4.1 | 1.0 ± 5.5 | 0.083 | 0.834 |
| Alcohol, % total energy intake | 2.5 ± 3.6 | 2.9 ± 4.3 | 0.1 ± 1.0 | 0.499 | 0.235 |
| **Appetite traits** |  |  |  |  |  |
| TFEQ - restraint | 8.5 ± 3.7 | 12.4 ± 3.6 | **4.4 ± 5.0** | **0.873** | 0.022 |
| TFEQ - disinhibition | 9.3 ± 3.5 | 8.6 ± 3.3 | **-1.9 ± 2.7** | **-0.697** | 0.055 |
| TFEQ - hunger | 5.4 ± 2.8 | 5.3 ± 3.3 | -0.7 ± 3.7 | -0.350 | 0.297 |
| Power of Food Scale | 12.8 ± 3.3 | 12.1 ± 1.8 | **-2.0 ± 3.1** | **-0.659** | 0.067 |
| Food Cravings Inventory | 41.6 ± 11.6 | 39.1 ± 10.2 | **-6.4 ± 9.1** | **-0.706** | 0.053 |
| **Appetite states\*\*\*** |  |  |  |  |  |
| Hunger | 34.3 ± 10.9 | 33.9 ± 12.2 | -0.4 ± 9.2 | -0.043 | 0.876 |
| Fullness | 45.8 ± 11.3 | 45.0 ± 12.4 | -0.8 ± 9.0 | -0.089 | 0.744 |
| Prospective food consumption | 41.1 ± 13.2 | 41.6 ± 14.1 | 0.6 ± 8.5 | 0.071 | 0.790 |
| **Physical activity\*\*\*** |  |  |  |  |  |
| Steps, n/day | 7732 ± 2592 | 6274 ± 2417 | **-1458 ± 2401** | **-0.607** | 0.106 |
| Sedentary time, min/day | 710 ± 139 | 669 ± 127 | -41 ± 115 | -0.360 | 0.312 |
| Sedentary time, %waking | 76 ± 14 | 74 ± 13 | -2 ± 12 | -0.164 | 0.636 |
| Light PA, min/day | 41 ± 12 | 40 ± 16 | -1 ± 9 | -0.126 | 0.716 |
| Light PA %waking | 4 ± 1 | 4 ± 2 | 0 ± 1 | 0.045 | 0.897 |
| Moderate-to-vigorous PA, min/day | 61 ± 31 | 46 ± 22 | **-16 ± 27** | **-0.572** | 0.125 |
| Moderate-to-vigorous PA, %waking | 7 ± 4 | 5 ± 3 | -2 ± 3 | -0.467 | 0.199 |
| Bouted moderate-to-vigorous PA, min/day | 6 ± 6 | 10 ± 10 | 5 ± 11 | 0.416 | 0.248 |

Data presented as mean ± SD. Predicted RMR based on population-specific linear regression using body composition. P-values for all outcomes except appetite states are from paired samples t-test or Wilcoxon signed rank tests if differences violated normality assumptions. Data from appetite states were derived from a linear mixed model including time point (baseline, follow-up) as a fixed effect, participant as a random effect, and adjustments for the day of the study, the time of the assessment, and energy intake of the meal prior to the assessment
FFM: fat free mass; FM: fat mass; HbA1c: glycated hemoglobin; HDL: high-density lipoprotein; HOMA-IR: homeostatic model assessment for insulin resistance; LDL: low-density lipoprotein; PA: physical activity; RMR: resting metabolic rate. Large effects are highlighted with **bold** text.
\*n=11 at follow-up
\*\*n=7 with valid data at follow-up
\*\*\*n=9 with valid data at follow-up

In absolute terms, RMR decreased by 102 ± 170 kcal/day. RMR at follow-up (1747 ± 227 kcal/day) was similar to predicted when accounting for body composition (1731 ± 242 kcal/day) and RMR residuals were modest (−20 ± 157 kcal/day).

#### 3.4.2 Behavioral Outcomes

Measures of appetite traits improved: dietary restraint increased while disinhibition, appetite for palatable foods, and food cravings decreased. Self-reported energy and macronutrient intake parameters remained similar from baseline to follow-up, with a notable increase in the percent energy intake from alcohol (**Table 3**). Large effect sizes were observed for decreases in steps/day and minutes of MVPA/day. There were no differences in EMA measures of hunger, satiety, or prospective food consumption at baseline compared to follow-up, **Table 3**. Large effect sizes were observed for decreases in steps/day and minutes of MVPA/day.

## 4. Discussion

The findings of our study provide proof-of-concept that integrating a breath-based yoga program into a BWL intervention is feasible and acceptable and may support weight loss comparable to that typically observed after 3–4 months of BWL participation. In fact, weight loss was comparable to that seen in DRIFT, a similar BWL study conducted at our institution, which included a higher physical activity prescription than our study (300 min/week of moderate intensity aerobic activity vs 150 min/week moderate intensity activity in our study) and also provided a free fitness center membership. Metabolic health and appetite traits that theoretically support long-term health and weight loss maintenance were also improved. Collectively, the observed effects point to a potential additive effect of breath-based yoga when incorporated into BWL programs. While the proof-of-concept was successful, further intervention refinement followed by randomized pilot trial, multiphase optimization strategy trial [37] and/or large-scale randomized clinical trials is warranted to more rigorously test our findings and evaluate the long-term effectiveness of yoga as a complementary modality for facilitating and sustaining weight loss.

It was feasible to recruit a sample of adults with overweight or obesity who are otherwise metabolically healthy in a short timeframe and complete a large number of rigorous outcome measures in the sample. Satisfaction among enrolled participants was high; however, attrition was higher than anticipated, suggesting improvements are needed. The uncertainty of the COVID-19 omicron surge and need to move classes temporarily online may have also contributed to the high attrition observed. Success in BWL interventions is highly related to completion of program requirements, such as attendance to in-person sessions and meeting diet and PA goals [38]; thus, it is crucial to characterize adherence and acceptability of novel interventions. In the present study, adherence to in-person session attendance and PA targets was similar [39] or superior [40,41] to previous BWL studies of similar duration (3-4 months), even despite the fact that part of the intervention occurred over the holidays and during the COVID-19 omicron surge. It is possible that being part of a group enhanced adherence, as feelings of belonging to a group and social connection can improve adherence [42]. Similar to previous research [43–45], participants in the present study reported satisfaction with the program in terms of the yoga component of the intervention itself and perceived positive changes in body weight, physical function, fatigue, psychological effects, overall well-being, and dietary intake as a result of the intervention. Thus, breath-based yoga may be a viable option to enhance and complement weight loss efforts for some individuals.

Despite the intervention substituting half of the aerobic exercise targets recommended by obesity treatment for weight loss (150 min/week vs. 300 min/week, with a sedentary to low-intensity yoga practice), we observed several positive changes in anthropometric and metabolic outcomes that reduce chronic disease risk. Weight loss was similar to a study using a higher PA target. Several participants in the present study achieved clinically meaningful weight loss, which is comparable to previous research with yoga practices focused on movement rather than breath. For example, a 2013 narrative review concluded that yoga can be effective for promoting weight loss and improving body composition, but more high-quality research including larger samples of individuals with overweight or obesity was needed [46]. This need was echoed in a 2016 systematic review and meta-analysis which reported that yoga effectively reduced BMI in the five studies that specifically included adults with overweight/obesity or metabolic syndrome (n=60 randomized to a yoga intervention in total pooled sample size) [6]. A more recent study examined the feasibility of a 6-month BWL intervention with either “restorative” or “vinyasa”-style yoga in 50 adults with obesity. Weight loss was approximately 4% at six months in both groups despite differences in the intensity of these *asana-*based yoga approaches. Such findings are expected because yoga *asanas –* whether “restorative” or “vinyasa”-style - are primarily classified as light intensity PA [47] and would not offer an equivalent replacement for energy expenditure from MVPA recommended for weight loss in current guidelines [3,48]. Collectively, these data underscore the possibility that factors beyond exercise volume and intensity may contribute to the weight loss observed when yoga and BWL supporting diet and PA modifications are combined.

Weight loss in the present study was accompanied by FFM loss without an excessive reduction in RMR. Any degree of weight loss is likely to be accompanied by FFM loss [49]. It is also worth noting that our intervention did not encourage resistance exercise training, which is known to help preserve FFM during weight loss efforts [50]. Future research should explore the feasibility and effects of incorporating resistance exercise with yoga in BWL interventions, especially in those at heightened risk for FFM loss (e.g., older adults).

Several recent systematic reviews and meta-analyses suggest yoga interventions are promising complementary approaches for improving metabolic health parameters such as glycemic control, lipid levels, and body composition among adults at-risk for or with type 2 diabetes [12,13,51–53]. Similarly, a previous six-month viniyoga (A ‘Hatha’ or movement-focused style of yoga) intervention found significant reductions in waist circumference compared to a waitlist control condition among post-treatment breast cancer survivors with overweight or obesity [54]. In line with these data, we observed trends in improved cardiometabolic health indicators (i.e., blood pressure, insulin, glucose, lipids, and waist circumference). These improvements are likely attributable to weight loss, with yoga potentially offering additional benefits in enhancing reductions in these biomarkers. Both obesity and chronic stress cause systemic inflammation, which is consistently and strongly associated with greater risk of morbidity and mortality [13,55]. Yoga has been shown to reduce measures of psychological and a physiological stress [11,56] and may decrease inflammation by attenuating reactivity of the hypothalamic-pituitary-adrenal axis and sympathoadrenal system. More rigorous studies that include measurements of these mediational inflammatory pathways are needed. The improvements in metabolic health indices following this breath-based yoga and BWL intervention are promising, particularly the reduction observed in systolic blood pressure. A 2017 meta-analysis of six yoga interventions showed a significant reduction in systolic blood pressure (mean difference: −4.59 mm Hg, 95% CI: −5.54 to −3.64) [57], nearly half of the mean difference in systolic blood pressure observed in the current study. These and other biomarker changes are important given the increasing prevalence of metabolic syndrome and its impacts on chronic disease risk [58,59]. These findings suggest that combining breath-based yoga with BWL could be especially beneficial for individuals with hypertension, offering an additional non-pharmacological approach to managing blood pressure.

Yoga may support weight loss by directly and indirectly affecting dietary intake and PA. Observational studies have found that regular yoga practice is associated with consuming more servings of fruits and vegetables, fewer sugar-sweetened beverages and snack foods, less frequent fast food consumption, and improved management of emotional eating [60]. However, our data and others [8] have not found decreases in energy intake in response to yoga, which may partly be due to the reliance on self-reported energy and macronutrient intake data. Appetite states and traits can provide insight into the more salient processes that underpin dietary intake. Ten minutes of slowed breathing (similar to some yoga breathing techniques) reduced hunger in females without obesity under experimental conditions designed to elicit food cravings [61]. Twice-weekly yoga after a standard BWL intervention resulted in less dietary lapses and continued weight loss compared to a control condition (cooking class) [62]. We found no differences in appetite states of hunger, satiety, and prospective food consumption. Interpretation is challenging, as some increase in hunger would be expected given the observed weight loss; in the absence of a control group, it remains unclear whether the intervention produced beneficial effects beyond those attributable to weight loss alone. The observed changes in appetite traits are in line with previous data that showed substantial improvements in dietary restraint, diet self-efficacy, and food cravings – all of which have been associated with weight loss [63,64]. Increased dietary restraint was particularly pronounced, and on par with or greater than seen in traditional BWL interventions [64–66]. Thus, our observation of unaltered dietary intake may stem from recognized systemic inaccuracies in self-reported data, but favorable modifications in dietary behaviors or traits in response to yoga may be indicative for lasting weight management. Yoga could serve as a potentially beneficial adjunct or alternative to approaches such as anti-obesity medications or bariatric surgery, given its effects on constructs like power of food and food cravings - effects that may parallel, albeit to a lesser degree, the reductions in food-related preoccupation commonly reported with anti-obesity medications [67].

As reported in a systematic review in people with overweight or obesity, yoga may increase PA [8]. However, the only study to measure objective (rather that subjective self-reported) PA during a yoga intervention in people with overweight or obesity did not find changes in MVPA, despite greater self-reported PA parameters [44]. Our finding of reduced steps and MVPA (in minutes/day) is perplexing but may be partially attributed to the high level of PA observed at baseline in some participants. Many gyms were closed or had limited capacity due to COVID-19 restrictions during the intervention (especially during the omicron surge in weeks 12 -14), which may have also contributed to reduced steps and MVPA. It is also possible that the time required to complete the yoga sessions displaced time for other physical activities. Despite null changes in accelerometer-based measures of PA, participants reported a reduction in perceived exercise barriers – a construct which has been associated with greater PA in previous research [68]. Yoga could mitigate perceived exercise barriers by reducing back and joint pain and improving physical function, balance, isometric strength, and cardiorespiratory fitness [60,69,70].

To our knowledge, this is the first study to assess the feasibility and preliminary effects of a breath-based yoga program embedded within a traditional BWL intervention on several parameters that are critical for weight loss in people with overweight or obesity. However, some limitations should be considered in the interpretation of our results. The sample size was limited and we did not include a control group, although the purpose of the study (i.e., feasibility) negated the need for a larger sample size or control group (in line with ORBIT guidelines [16]). As this was a single-arm trial, isolating the specific effects of yoga from those of weight loss would require larger, multi-arm trials designed to disentangle these components. Furthermore, the sample was composed of mostly white, middle-aged, highly educated females, which may limit the generalizability of our findings.

In conclusion, a combined breath-based yoga and BWL intervention is highly acceptable, results in weight loss, and positively impacts several parameters that can contribute to negative energy balance and reduce chronic disease risk. Continued exploration of the role of yoga on initial and sustained weight loss in people with overweight or obesity is warranted.

## Author contributions

**SAP** contributed to acquiring data, conducting data analyses, and wrote the first draft of the manuscript. **PD** contributed to developing the yoga intervention. **VC** contributed to conceiving and refining the study. **KB** contributed to creating and leading the behavioral weight loss intervention. **KF** contributed to data analyses. **AEC** conceived the study, contributed to data collection and analyses, and acquired funding for the study. All authors contributed to revising the manuscript and approved the final version.

## Data Availability

All data produced in the present study are available upon reasonable request to the authors.

## References

1. Hales CM, Fryar CD, Carroll MD, Freedman DS, Ogden CL. Trends in Obesity and Severe Obesity Prevalence in US Youth and Adults by Sex and Age, 2007-2008 to 2015-2016. JAMA. 2018;319(16):1723. doi:10.1001/jama.2018.3060

2. Cawley J, Biener A, Meyerhoefer C, et al. Direct medical costs of obesity in the United States and the most populous states. JMCP. 2021;27(3):354–366. doi:10.18553/jmcp.2021.20410

3. Jensen MD, Ryan DH, Apovian CM, et al. 2013 AHA/ACC/TOS guideline for the management of overweight and obesity in adults: a report of the American College of Cardiology/American Heart Association Task Force on Practice Guidelines and The Obesity Society. Circulation. 2014;129(25 Suppl 2):S102–38. doi:10.1161/01.cir.0000437739.71477.ee

4. Anderson JW, Konz EC, Frederich RC, Wood CL. Long-term weight-loss maintenance: a meta-analysis of US studies. Am J Clin Nutr. 2001;74(5):579–584.

5. Ramamoorthi R, Gahreman D, Skinner T, Moss S. The effect of yoga practice on glycemic control and other health parameters in the prediabetic state: A systematic review and meta-analysis. Abbasalizad Farhangi M, ed. PLoS ONE. 2019;14(10):e0221067. doi:10.1371/journal.pone.0221067

6. Lauche R, Langhorst J, Lee MS, Dobos G, Cramer H. A systematic review and meta-analysis on the effects of yoga on weight-related outcomes. Preventive medicine. 2016;87:213–232. doi:10.1016/j.ypmed.2016.03.013

7. Chu P, Gotink RA, Yeh GY, Goldie SJ, Hunink MM. The effectiveness of yoga in modifying risk factors for cardiovascular disease and metabolic syndrome: A systematic review and meta-analysis of randomized controlled trials. Eur J Prev Cardiolog. 2016;23(3):291–307. doi:10.1177/2047487314562741

8. Caldwell AE, Purcell SA, Gray B, Smieja H, Catenacci VA. The impact of yoga on components of energy balance in adults with overweight or obesity: A systematic review. Obesity Science & Practice. 2022;8(2):219–232. doi:10.1002/osp4.552

9. Gard T, Noggle JJ, Park CL, Vago DR, Wilson A. Potential self-regulatory mechanisms of yoga for psychological health. Front Hum Neurosci. 2014;8:770. doi:10.3389/fnhum.2014.00770

10. Field T. Yoga research review. Complementary therapies in clinical practice. 2016;24:145–161. doi:10.1016/j.ctcp.2016.06.005

11. Pascoe MC, Bauer IE. A systematic review of randomised control trials on the effects of yoga on stress measures and mood. Journal of Psychiatric Research. 2015;68:270–282. doi:10.1016/j.jpsychires.2015.07.013

12. Thind H, Lantini R, Balletto BL, et al. The effects of yoga among adults with type 2 diabetes: A systematic review and meta-analysis. Preventive Medicine. 2017;105:116–126. doi:10.1016/j.ypmed.2017.08.017

13. Innes KE, Selfe TK. Yoga for Adults with Type 2 Diabetes: A Systematic Review of Controlled Trials. Journal of Diabetes Research. 2016;2016:1–23. doi:10.1155/2016/6979370

14. Jakicic JM, Davis KK, Rogers RJ, et al. Feasibility of Integration of Yoga in a Behavioral Weight-Loss Intervention: A Randomized Trial. Obesity. 2021;29(3):512–520. 10.1002/oby.23089

15. Powell LH, Appelhans BM, Ventrelle J, et al. Development of a lifestyle intervention for the metabolic syndrome: Discovery through proof-of-concept. Health Psychology. 2018;37(10):929–939. doi:10.1037/hea0000665

16. Czajkowski SM, Powell LH, Adler N, et al. From ideas to efficacy: The ORBIT model for developing behavioral treatments for chronic diseases. Health Psychology. 2015;34(10):971–982. doi:10.1037/hea0000161

17. Ostendorf DM, Caldwell AE, Zaman A, et al. Comparison of weight loss induced by daily caloric restriction versus intermittent fasting (DRIFT) in individuals with obesity: study protocol for a 52-week randomized clinical trial. Trials. 2022;23(1):718. doi:10.1186/s13063-022-06523-2

18. Peters JC, Wyatt HR, Foster GD, et al. The effects of water and non-nutritive sweetened beverages on weight loss during a 12-week weight loss treatment program. Obesity (Silver Spring). 2014;22(6):1415–1421. doi:10.1002/oby.20737

19. Wyatt HR, Jortberg BT, Babbel C, et al. Weight loss in a community initiative that promotes decreased energy intake and increased physical activity and dairy consumption: Calcium Weighs-In. Journal of physical activity & health. 2008;5(1):28–44. doi:10.1123/jpah.5.1.28

20. Human Energy Requirements: Report of a Joint FAO/WHO/UNU Expert Consultation. Food and Agriculture Organization, FAO Food and Nutrition Technical Report Series No. 1; 2004.

21. The Diabetes Prevention Program. Design and methods for a clinical trial in the prevention of type 2 diabetes. Diabetes Care. 1999;22(4):623–634.

22. American College of Sports Medicine, Riebe D, Ehrman JK, Liguori G, Magal M. ACSM’s Guidelines for Exercise Testing and Prescription. Tenth edition. Wolters Kluwer; 2018.

23. Services USD of H and H. Physical Activity Guidelines for Americans. 2nd ed. 2018;(APril 6, 2020).

24. Catenacci VA, Ostendorf DM, Pan Z, et al. The Effect of 4:3 Intermittent Fasting on Weight Loss at 12 Months : A Randomized Clinical Trial. Ann Intern Med. Published online April 1, 2025. doi:10.7326/ANNALS-24-01631

25. Williamson DA, Bray GA, Ryan DH. Is 5% weight loss a satisfactory criterion to define clinically significant weight loss? Obesity. 2015;23(12):2319–2320. doi:10.1002/oby.21358

26. Burrows TL, Ho YY, Rollo ME, Collins CE. Validity of Dietary Assessment Methods When Compared to the Method of Doubly Labeled Water: A Systematic Review in Adults. Vol 10.; 2019. https://www.frontiersin.org/articles/10.3389/fendo.2019.00850

27. Organization WH. Obesity: Preventing and Managing the Global Epidemic. Report of a WHO Consultation. Vol 894.; 2000:1–253.

28. Tai MM. A Mathematical Model for the Determination of Total Area Under Glucose Tolerance and Other Metabolic Curves. Diabetes Care. 1994;17(2):152–154. doi:10.2337/diacare.17.2.152

29. Weir JB. New methods for calculating metabolic rate with special reference to protein metabolism. 1949. Nutrition. 1990;6:213–221.

30. Fothergill E, Guo J, Howard L, et al. Persistent metabolic adaptation 6 years after “The Biggest Loser” competition. Obesity (Silver Spring). 2016;24:1612–1619. doi:10.1002/oby.21538

31. Wolfe BM, Schoeller DA, McCrady-Spitzer SK, Thomas DM, Sorenson CE, Levine JA. Resting Metabolic Rate, Total Daily Energy Expenditure, and Metabolic Adaptation 6 Months and 24 Months After Bariatric Surgery. Obesity (Silver Spring, Md). 2018;26(5):862–868. doi:10.1002/oby.22138

32. Galgani JE, Santos JL. Insights about weight loss-induced metabolic adaptation. Obesity. 2016;24(2):277–278. 10.1002/oby.21408

33. McCrory MA, McCrory MA, Hajduk CL, Roberts SB. Procedures for screening out inaccurate reports of dietary energy intake. Public Health Nutr. 2002;5(6A):873–882. doi:10.1079/PHN2002387

34. Stunkard AJ, Messick S. The three-factor eating questionnaire to measure dietary restraint, disinhibition and hunger. Journal of psychosomatic research. 1985;29(1):71–83. doi:10.1016/0022-3999(85)90010-8

35. Lowe MR, Butryn ML, Didie ER, et al. The Power of Food Scale. A new measure of the psychological influence of the food environment. Appetite. 2009;53(1):114–118. doi:10.1016/j.appet.2009.05.016

36. White MA, Whisenhunt BL, Williamson DA, Greenway FL, Netemeyer RG. Development and validation of the food-craving inventory. Obesity research. 2002;10(2):107–114. doi:10.1038/oby.2002.17

37. Collins LM, Murphy SA, Strecher V. The multiphase optimization strategy (MOST) and the sequential multiple assignment randomized trial (SMART): new methods for more potent eHealth interventions. Am J Prev Med. 2007;32(5 Suppl):S112–118. doi:10.1016/j.amepre.2007.01.022

38. Acharya SD, Elci OU, Sereika SM, et al. Adherence to a behavioral weight loss treatment program enhances weight loss and improvements in biomarkers. Patient Prefer Adherence. 2009;3:151–160.

39. Travier N, Fonseca-Nunes A, Javierre C, et al. Effect of a diet and physical activity intervention on body weight and nutritional patterns in overweight and obese breast cancer survivors. Med Oncol. 2014;31(1):783. doi:10.1007/s12032-013-0783-5

40. Befort CA, Nollen N, Ellerbeck EF, Sullivan DK, Thomas JL, Ahluwalia JS. Motivational interviewing fails to improve outcomes of a behavioral weight loss program for obese African American women: a pilot randomized trial. J Behav Med. 2008;31(5):367–377. doi:10.1007/s10865-008-9161-8

41. Colley RC, Hills AP, O’Moore-Sullivan TM, Hickman IJ, Prins JB, Byrne NM. Variability in adherence to an unsupervised exercise prescription in obese women. Int J Obes (Lond). 2008;32(5):837–844. doi:0803799 [pii] 10.1038/sj.ijo.0803799

42. Rogers M, Lemstra M, Bird Y, Nwankwo C, Moraros J. Weight-loss intervention adherence and factors promoting adherence: a meta-analysis. PPA. 2016;Volume 10:1547–1559. doi:10.2147/PPA.S103649

43. Cohen BE, Chang AA, Grady D, Kanaya AM. Restorative yoga in adults with metabolic syndrome: a randomized, controlled pilot trial. Metabolic syndrome and related disorders. 2008;6(3):223–229. doi:10.1089/met.2008.0016

44. Mama SK, Bhuiyan N, Chaoul A, et al. Feasibility and acceptability of a faith-based mind-body intervention among African American adults. Translational behavioral medicine. 2020;10(4):928–937. doi:10.1093/tbm/iby114

45. Ruby M, Repka CP, Arciero PJ. Comparison of Protein-Pacing Alone or With Yoga/Stretching and Resistance Training on Glycemia, Total and Regional Body Composition, and Aerobic Fitness in Overweight Women. Journal of physical activity & health. 2016;13(7):754–764. doi:10.1123/jpah.2015-0493

46. Rioux JG, Ritenbaugh C. Narrative review of yoga intervention clinical trials including weight-related outcomes. Altern Ther Health Med. 2013;19(3):32–46.

47. Larson-Meyer DE. A Systematic Review of the Energy Cost and Metabolic Intensity of Yoga. Med Sci Sports Exerc. 2016;48(8):1558–1569. doi:10.1249/MSS.0000000000000922

48. Donnelly JE, Blair SN, Jakicic JM, Manore MM, Rankin JW, Smith BK. American College of Sports Medicine Position Stand. Appropriate physical activity intervention strategies for weight loss and prevention of weight regain for adults. Medicine and science in sports and exercise. 2009;41(2):459–471. doi:10.1249/MSS.0b013e3181949333

49. Chaston TB, Dixon JB, O’Brien PE. Changes in fat-free mass during significant weight loss: a systematic review. International Journal of Obesity. 2007;31(5):743–750. doi:10.1038/sj.ijo.0803483

50. Heymsfield SB, Gonzalez MCC, Shen W, Redman L, Thomas D. Weight loss composition is one-fourth fat-free mass: a critical review and critique of this widely cited rule. Obes Rev. 2014;15(4):310–321. doi:10.1111/obr.12143

51. Cui J, Yan J, Yan L, Pan L, Le J, Guo Y. Effects of yoga in adults with type 2 diabetes mellitus: A meta-analysis. J of Diabetes Invest. 2017;8(2):201–209. doi:10.1111/jdi.12548

52. Kumar V, Jagannathan A, Philip M, Thulasi A, Angadi P, Raghuram N. Role of yoga for patients with type II diabetes mellitus: A systematic review and meta-analysis. Complementary Therapies in Medicine. 2016;25:104–112. doi:10.1016/j.ctim.2016.02.001

53. Jayawardena R, Ranasinghe P, Chathuranga T, Atapattu PM, Misra A. The benefits of yoga practice compared to physical exercise in the management of type 2 Diabetes Mellitus: A systematic review and meta-analysis. Diabetes & Metabolic Syndrome: Clinical Research & Reviews. 2018;12(5):795–805. doi:10.1016/j.dsx.2018.04.008

54. Littman AJ, Bertram LC, Ceballos R, et al. Randomized controlled pilot trial of yoga in overweight and obese breast cancer survivors: effects on quality of life and anthropometric measures. Supportive care in cancer : official journal of the Multinational Association of Supportive Care in Cancer. 2012;20(2):267–277. doi:10.1007/s00520-010-1066-8

55. Cohen S, Janicki-Deverts D, Miller GE. Psychological Stress and Disease. JAMA. 2007;298(14):1685. doi:10.1001/jama.298.14.1685

56. Riley KE, Park CL. How does yoga reduce stress? A systematic review of mechanisms of change and guide to future inquiry. Health Psychol Rev. 2015;9(3):379–396. doi:10.1080/17437199.2014.981778

57. Park SH, Han KS. Blood Pressure Response to Meditation and Yoga: A Systematic Review and Meta-Analysis. The Journal of Alternative and Complementary Medicine. 2017;23(9):685–695. doi:10.1089/acm.2016.0234

58. Liang X, Or B, Tsoi MF, Cheung CL, Cheung BMY. Prevalence of metabolic syndrome in the United States National Health and Nutrition Examination Survey 2011–18. Postgraduate Medical Journal. 2023;99(1175):985–992. doi:10.1093/postmj/qgad008

59. Alberti KGMM, Zimmet P, Shaw J. The metabolic syndrome--a new worldwide definition. Lancet. 2005;366(9491):1059–1062. doi:10.1016/S0140-6736(05)67402-8

60. Watts AW, Rydell SA, Eisenberg ME, Laska MN, Neumark-Sztainer D. Yoga’s potential for promoting healthy eating and physical activity behaviors among young adults: a mixed-methods study. The international journal of behavioral nutrition and physical activity. 2018;15(1):42. doi:10.1186/s12966-018-0674-4

61. Meule A, Kübler A. A Pilot Study on the Effects of Slow Paced Breathing on Current Food Craving. Applied psychophysiology and biofeedback. 2017;42(1):59–68. doi:10.1007/s10484-017-9351-7

62. Unick JL, Dunsiger SI, Bock BC, et al. A randomized trial examining the effect of yoga on dietary lapses and lapse triggers following behavioral weight loss treatment. Obesity Science & Practice. 2023;9(5):484–492. doi:10.1002/osp4.678

63. Bates S, Norman P, Breeze P, Brennan A, Ahern AL. Mechanisms of Action in a Behavioral Weight-Management Program: Latent Growth Curve Analysis. Annals of Behavioral Medicine. 2022;56(1):64–77. doi:10.1093/abm/kaab019

64. Papini NM, Foster RNS, Lopez NV, Ptomey LT, Herrmann SD, Donnelly JE. Examination of three-factor eating questionnaire subscale scores on weight loss and weight loss maintenance in a clinical intervention. BMC Psychol. 2022;10(1):101. doi:10.1186/s40359-022-00806-8

65. Jakicic JM, Rogers RJ. Association of Eating Behaviors with Variability in Weight Change in Response to Physical Activity Interventions in Adults with Overweight. Nutrients. 2023;15(15):3452. doi:10.3390/nu15153452

66. Purcell SA, Legget KT, Halliday TM, et al. Appetitive and Metabolic Responses to an Exercise versus Dietary Intervention in Adults with Obesity. Translational Journal of the American College of Sports Medicine. 2022;7(4). https://journals.lww.com/acsm-tj/fulltext/2022/10140/appetitive_and_metabolic_responses_to_an_exercise.10.aspx

67. Hayashi D, Edwards C, Emond JA, et al. What Is Food Noise? A Conceptual Model of Food Cue Reactivity. Nutrients. 2023;15(22). doi:10.3390/nu15224809

68. Call CC, Roberts SR, Schumacher LM, Remmert JE, Kerrigan SG, Butryn ML. Perceived barriers to physical activity during and after a behavioural weight loss programme. Obesity Science & Practice. 2020;6(1):10–18. doi:10.1002/osp4.373

69. Alert MD, Rastegar S, Foret M, et al. The effectiveness of a comprehensive mind body weight loss intervention for overweight and obese adults: a pilot study. Complementary therapies in medicine. 2013;21(4):286–293. doi:10.1016/j.ctim.2013.05.005

70. Villiot-Danger JC, Villiot-Danger E, Borel JC, Pépin JL, Wuyam B, Vergès S. Respiratory muscle endurance training in obese patients. Int J Obes. 2011;35(5):692–699. doi:10.1038/ijo.2010.191

