## Supplemental Table 1 for "Complementing Behavioral Weight Loss with Breath-based Yoga: A Proof-of-Concept and Initial Feasibility Study"

**Supplemental Table 1. Self-reported changes in health and wellbeing during the yoga intervention**

|  | **Week 8** | **Week 16** |
| --- | --- | --- |
|  | **n (%) ‘yes’** | **n (%) ‘yes’** |
| Are there any physical effects (i.e., fatigue, strength, endurance, hunger, sleep quality/quantity, increased mobility) that you feel changed as a result of the yoga and/or diet and physical activity program? | 8 (72.7) | 10 (90.9) |
| Are there any psychological effects (i.e., stress, confidence, body satisfaction) that you feel changed as a result of the yoga and or diet and physical activity program? | 7 (63.6) | 10 (90.9) |
| Do you think your overall well-being changed as a result of in the yoga and/or diet and physical activity program? | 10 (90.9) | 10 (90.9) |
| Do you feel like your diet changed during this study? | 10 (90.9) | 10 (90.9) |
